# Nursing and midwifery students’ perceptions and experiences of clinical learning in Papua New Guinea

**DOI:** 10.1101/2025.06.19.25329826

**Authors:** Nikki Naki Joseph, Gracelyn Potjepat, Marian Vowari Minnala, Paula Zebedee Aines, Lilian Temo, McKenzie Maviso

## Abstract

**Introduction:** Clinical learning environment is essential for developing the skills and competence of nursing and midwifery students; however, it can affect their ability to apply theory into practice. This study aimed to explore the perceptions and experiences of student nurses and midwives regarding their clinical learning in Papua New Guinea (PNG).

**Methods:** A phenomenological study was conducted using purposive sampling to recruit 18 nursing and midwifery students. Data were collected through individual, in-depth semi-structured interviews and analyzed using thematic analysis.

**Results:** Four themes emerged from the interview data, influencing participants’ perceptions and experiences regarding clinical learning: (1) supportive clinical environment, (2) perceived bias and dismissive attitudes, (3) poor supervision and unmet learning outcomes, and (4) clinical resource constraints.

**Conclusions:** Findings revealed that negative clinician attitudes, poor supervision, and limited resources hindered students’ clinical learning. Strengthening collaboration between institutions and hospitals, along with improved resource support, is essential for better learning outcomes.

## Introduction

Clinical learning and practice constitute an indispensable component of nursing education, where students integrate cognitive, affective, and psychomotor skills to develop critical clinical competencies for practice (Jacob, Seif and Munyaw, 2023). Using the nursing competency scale, Meretoja et al. (2004) described clinical competence as the ability of students to integrate knowledge, skills, attitudes, and values into nursing practices, which remain important for professional standards. Nursing and midwifery students must apply their theoretical knowledge as they confront challenges that help build their courage and confidence in real-life clinical situations (Aryuwat *et al*., 2024). Clinical practice challenges and skill acquisition foster critical thinking, decision-making, and emotional resilience, enhancing adaptability and professional confidence for a seamless career transition (Jacob, Seif and Munyaw, 2023).

The clinical learning environment comprises the dynamic factors within clinical settings where students apply theory, develop skills, and enhance problem-solving and reasoning abilities (Wuni *et al*., 2025). A typical clinical learning environment includes health professionals, nurse educators, and patients, which affects nursing students’ careers either positively or negatively by impacting their performance (Jacob, Seif and Munyaw, 2023). Four distinct attributes of a clinical learning environment are the physical environment, interpersonal and psychosocial aspects, organizational culture, and clinical teaching components (Flott and Linden, 2016). During clinical placements, the combination of practical skills, theoretical knowledge, and quality mentorship significantly bolster nursing students’ motivation, confidence, and commitment to the profession (Wuni *et al*., 2025).

Given the significance of clinical learning, a growing body of evidence highlights a strong correlation between the quality of the learning environment and nursing students’ satisfaction (Cant, Ryan and Cooper, 2021). A supportive clinical learning environment that encompasses effective supervision, constructive feedback, and a collaborative culture, enhances coping abilities, improves learning opportunities, and supports the growth of skilled and resilient practitioners (Jacob, Seif and Munyaw, 2023). A study conducted in Spain discovered that nursing students expressed high levels of satisfaction with their clinical learning environment and supervision at hospitals where they completed clinical placements, suggesting a positive relationship between these elements (Rodríguez-García *et al*., 2021). A strong sense of belonging during clinical placements can boost students’ confidence and enhance their motivation to learn. As Mikkonen et al. (2020) highlighted, clinical mentoring is crucial for the professional development of nursing students and can significantly increase their motivation to enter and remain in the profession.

Students may struggle to apply theoretical knowledge in practice when the clinical learning environment lacks sufficient support or fails to foster a positive atmosphere. Research has shown that negative experiences and poor learning environments, marked by negative attitudes from staff toward working with seniors, adversely impact students’ learning (Zhang *et al*., 2022). Students’ clinical learning can also be influenced by various interpersonal, sociocultural, instructional, environmental, emotional, and physical factors, including poor teaching materials, strained professional relationships, inadequate supervision, limited clinical resources, and a hostile work environment (Amoo *et al*., 2022).

Papua New Guinea (PNG) has one of the world’s most dispersed and isolated populations, characterized by its diverse cultures. The country has a decentralized healthcare system based on primary healthcare principles (Grundy *et al*., 2019). Access to healthcare services is variable as it is often affected by poor health infrastructure, and limited transport access (Grundy *et al*., 2019). In addition, persistent health workforce shortages in PNG continue to be driven by chronic underinvestment, poor working conditions, and imbalances between workforce supply and demand (Dimiri *et al*., 2022; Renton and Muddle, 2024). There is considerable demand for a strengthened health workforce and equitable healthcare in PNG, with existing disparities highlighting the urgent need for expanded health education and training.

Previous studies on nursing and midwifery education in PNG focused on the challenges new graduate midwives face and the broad aspects of nursing and midwifery education and regulation (Moores *et al*., 2016; Goveh *et al*., 2022). There are currently no published studies that specifically examine the perceptions and experiences of nursing or midwifery students in their clinical learning environments in PNG. Additionally, there is a lack of literature focused on nursing and midwifery education in the country. Addressing this knowledge gap is crucial for enhancing the quality of nursing and midwifery education, improving clinical learning practices. Therefore, this study aims to explore nursing and midwifery students’ perceptions and experiences regarding their clinical learning practices.

## Methods

### Study design

Informed by an interpretive philosophical paradigm, this study employed a hermeneutic phenomenological approach informed by van Manen (2016) to explore the lived experiences of nursing and midwifery students. This method emphasizes interpreting personal experiences by providing phenomenological descriptions that reveal the meaning and significance of the phenomena as experienced by participants (Laverty, 2003). The hermeneutical aspect involves interpreting these descriptions based on the principle of understanding the experiences shared by participants (Laverty, 2003). Qualitative research aims to explore lived experiences, emphasizing trust, transparency, and flexibility, making it well-suited for insights in nursing and midwifery education (Polit and Beck, 2008).

### Sampling

Purposive sampling was used to select 18 students from the Bachelor of Clinical Nursing programs at the School of Medicine and Health Sciences, University of Papua New Guinea (UPNG): Child Health (n=3), Midwifery (n=6), Mental Health (n=2), and Critical Care (n=7). Purposive sampling is a type of non-probability sampling method that involves researchers selecting a specific group of individuals based on characteristics that are relevant to the study (Stratton, 2023). This approach was appropriate as it allowed for the collection of rich insights into the clinical learning experiences of nursing and midwifery students. Students from other health disciplines were excluded from this study.

### Data Collection

Following informed consent, the primary researchers used a semi-structured interview guide consisting of four main questions to guide the discussion. Participants were asked, ‘Please, tell us about your clinical experiences at the hospital. Tell us more about the…’ Follow-up questions were posed based on their statements and responses. Additionally, probing questions were asked regarding the participants’ responses and opinions (e.g., “Would you elaborate more on this?” or “What did you mean by saying…?”) to obtain in-depth information. The interviews provided participants with a platform to lead discussions and share their perceptions and experiences regarding clinical learning. In alignment with hermeneutic phenomenology (van Manen, 2016), interviews fostered the development of conversational relationships with participants, allowing for a deeper exploration and reflection on the significance of their experiences. All interviews were recorded and transcribed verbatim after the interview sessions. Each interview lasted approximately 30 to 50 minutes. The interview process continued until data saturation was achieved. To ensure credibility, audio recordings and transcripts were consistently reviewed for alignment with data interpretation, identified codes, and emerging themes. All interviews were audio-recorded and transcribed verbatim.

### Data Analysis

Using hermeneutic phenomenological data analysis techniques, recurring themes were identified, leading to a ‘thick description’ of participants’ experiences (van Manen, 2016). A thematic analysis was conducted using a six-step process to examine qualitative data, systematically identifying and organizing patterns of meaning into themes that provide deeper insights into participants’ perspectives and experiences (Braun and Clarke, 2022). To gain familiarity with the data, transcripts were systematically and openly read. Non-verbal ques such as pauses, laughter, vocalizations, and facial expressions were removed, and grammar was corrected to improve readability without altering the meaning of participants’ narratives. Data were manually analyzed by organizing notes, codes, and categories in Microsoft Word, based on shared meanings. These were then examined for patterns, leading to the development of sub-themes and overarching themes. The themes were further refined and selected to accurately reflect participants’ perceptions and experiences (Braun and Clarke, 2022). An independent reviewer (not involved in the study) examined the themes for validation. Reporting adheres to the Standards for Reporting Qualitative Research (SRQR) Checklist (O’Brien *et al*., 2014).

### Rigor

This study used four criteria to establish rigor: (1) credibility, (2) transferability, (3) dependability, and (4) confirmability (Cypress, 2017). First, credibility was established based on several strategies throughout the study. During data collection, participants were actively engaged in interviews, followed by debriefing sessions to review the process. Four independent researchers were involved in the data analysis and coding of the themes. The researchers reviewed each theme to ensure that it accurately reflected the participants’ narratives. Second, the transferability of the findings was ensured by providing a detailed description of the study context. Third, dependability was established through the dense description of the methodology used and the description of the data. All interview materials, transcriptions, findings, interpretations, and recommendations were kept accessible to the principal investigator and supervisor to allow for an audit trail; descriptions, codes, and themes were also confirmed. Finally, confirmability was enhanced by using verbatim quotations from participants’ narratives and incorporating field notes, which helped minimize researcher-induced biases (Cypress, 2017). The data were shared among colleagues for peer review and analysis to ensure trustworthiness.

### Ethical considerations

Ethical approval for this study was obtained from the Research Ethics Committee of the School of Medicine and Health Sciences, UPNG (02-07-2024). Students received information sheets and provided informed consent before participating in the study.

## Results

A total of 18 nursing and midwifery students (3 males and 15 females) participated in this study. Students were between the ages of 30 and 40 with more than five years of clinical experience in rural and urban healthcare facilities. Analysis of the data identified four overarching themes: (1) supportive clinical environment, (2) perceived bias and dismissive attitudes, (3) poor supervision and unmet learning outcomes, and (4) clinical resource constraints.

### Supportive Clinical Environment

Participants indicated that the clinical environment, which was friendly and supportive, enabled them to translate theoretical knowledge into clinical competence and effectively apply practice, enhancing the learning experience:

> “*When I first arrived at my assigned clinical workplace, I realized it was a completely new environment. I was confused and anxious, not knowing where essential items like the emergency trolley were located But the staff were friendly and helpful… they showed me where medical or emergency equipment is kept*… *I could work confidently*.*” (Participant 3)*
>
> *“The staff has been very supportive. They guided us through their workplace orientation and called us in whenever they needed to demonstrate a procedure… they also allowed us to perform some procedures. This helped us build our confidence as students*.*” (Participant 10)*

### Perceived Bias and Dismissive Attitudes

Students reported minimal support and dismissive attitudes from some clinical staff, with perceived bias during supervision and assessment contributing to dissatisfaction and reduced learning opportunities. Students also highlighted poor clinician engagement, especially when supervision was required. As one student shared:

> *“Some staff members were friendly, but I noticed that others were selective in their assistance when it came to assessing students’ clinical competencies. Also, some staff did not communicate well with us; they didn’t greet us and responded to our questions in an unfriendly manner*.*” (Participant 4)*
>
> Another student echoed this concern, highlighting the impact on their learning:
>
> *“Sometimes, we were not greeted by the staff in the ward; instead, they just ignored us, I do not know why this happened. Some would walk past us without acknowledging our presence… it was difficult to work with them*.*” (Participant 5)*

### Poor Supervision and Unmet Learning Outcomes

Students expressed dissatisfaction with the quality of clinical supervision, noting its impact on their skill development and competency. In many cases, supervision during clinical placements was inadequate, as one participant stated:

> *“We just do our clinical procedures on our own… there was no proper clinical supervision. Some nurses would tell us to do what we can with our procedures. After the procedure is completed, they just sign the clinical logbooks. This is not good!” (Participant 9)*

Some students reported having to perform clinical tasks without supervision, being instructed to call for assistance only when needed. This lack of consistent oversight made it difficult to connect theoretical knowledge with practical skills. As one student explained:

> *“There is no (clinical) supervision…the nurses are just instructed to perform clinical procedures independently. If you feel confused or encounter an emergency, please call us for assistance. That was their advice to us…” (Participant 2)*

### Clinical Resource Constraints

Students consistently reported challenges related to the availability and use of clinical resources during placements, alongside concerns about poor infection control practices. Limited access to essential equipment not only hindered learning but also compromised safe practice standards. As two students explained:

> *“We do not have enough resources, including sterile equipment, such as suture trays and gloves to do a vaginal examination, especially in the Labor ward…so most of the time we run out and use unsterile tools/equipment to do our procedures*.*” (Participate 7)*
>
> *“We don’t have enough delivery trays in the labor ward. We wash and reuse them, sometimes just rinsing or using alcohol swabs. We’re not using sterile techniques*.*” (Participant 8)*

## Discussion

The study used a hermeneutic phenomenological approach to explore nursing and midwifery students’ perceptions and experiences of clinical learning. Findings from this study revealed mixed responses. While a supportive clinical environment contributed to positive learning experiences, students’ clinical learning was negatively affected by limited engagement, perceived bias and dismissive attitudes from clinicians, poor supervision and unmet learning objectives, and clinical resource constraints.

A supportive clinical environment promotes skill development, bridges theory and practice, and boosts student engagement and learning outcomes. In their study, Rodríguez-García *et al*. (2021) argued that the clinical learning environment directly impacts clinical performance and learning, serving as a vital link between academic instruction and practical skill development. Evidence indicates that a supportive clinical environment, underpinned by a positive workplace culture, enables nursing and midwifery students to actively engage in clinical learning, strengthen their clinical skills and competencies, and cultivate meaningful interpersonal and professional relationships (Rajjoub and AlMukhtar, 2024). Similarly, nursing and midwifery students gain confidence and demonstrate a greater willingness to learn when the clinical environment is supportive, characterized by open communication, constructive feedback, trust, and mutual respect between clinicians and students (Zhang *et al*., 2022). This study highlights how a supportive clinical learning environment improves student engagement, knowledge acquisition, clinical skills, and confidence.

In contrast, the study’s findings indicated that clinical nurses exhibited prejudice and negative attitudes while supervising nursing and midwifery students. Unethical attitudes and behaviors in healthcare can affect student learning, performance, satisfaction, and patient outcomes (Oshodi and Sookhoo, 2025). Recent studies confirm that clinician biases and unsupportive attitudes toward clinical supervision can negatively affect students’ clinical learning (Anagor *et al*., 2021). Negative attitudes from clinicians can hinder nursing and midwifery students’ learning by creating an unsupportive environment that limits their ability to ask questions, practice skills, and build confidence (Oshodi and Sookhoo, 2025). However, Jacob, Seif and Munyaw (2023) argued that A supportive clinical environment, combined with effective training, significantly enhances students’ confidence and improves their clinical learning experiences. Also, positive staff attitudes and mentorship encourage nursing and midwifery students to actively participate in clinical practice, build skills, and enhance their learning (Li *et al*., 2024). Future research could investigate the barriers to student learning that stem from nurses’ limited engagement and inadequate supervision in clinical settings.

Clinical supervisors play crucial roles in the development of students’ clinical competence and confidence, guaranteeing safe and patient-centered care. Students in our study claimed that a lack of clinical supervision was the most significant factor contributing to their dissatisfaction, a finding that reflects recent studies on clinical learning (Atashi *et al*., 2024). A possible reason for this issue is the lack of competency among clinical supervisors, who may not have the authority to effectively support nursing students during their placements. It can be difficult for nurses to provide effective supervision while also managing their daily patient care responsibilities (Tuomikoski *et al*., 2020). Another reason for inadequate clinical supervision could be the shortage of qualified supervisors and insufficient preceptorship trainings. This lack of oversight may stem from inadequate infrastructure and a lack of formal training, which can negatively impact students’ learning and practice (Atashi *et al*., 2024). Clinical preceptorship plays a vital role in building students’ confidence, competence, and professionalism, supporting their transition into nursing and midwifery practice (Walker and Norris, 2020; Atashi *et al*., 2024).

The study found that equipment and resource constraints during clinical placements hindered nursing and midwifery students from effectively applying theoretical knowledge in practice. The findings are in agreement with a similar study conducted in Tanzania (Jacob, Seif and Munyaw, 2023). Negative clinical experiences may contribute to the theory–practice gap, leading to reduced competence and confidence among nursing students upon graduation. Additionally, students in this study reported increased use of non-sterile techniques during clinical procedures, likely due to a lack of sterile equipment in clinical settings, as noted in a study from Malawi (Mbakaya *et al*., 2020). Limited clinical resources hinder effective participation in clinical practice, reducing the quality of learning experiences. Adequate availability and accessibility of medical resources and sterile equipment in clinical settings can improve students’ ability to engage effectively in clinical practice, which fosters self-’motivation, confidence, and competency (Gemuhay *et al*., 2019). The study emphasized the demand for health institutions to provide adequate resources and build the capacity to improve students’ learning in clinical settings.

This study explored nursing and midwifery students’ perceptions and experiences of clinical teaching and learning, which remain essential for enhancing clinical practice. While the findings align with existing research on clinical learning across various settings, there exist some study limitations. Firstly, it focused exclusively on students from a single institution, which may not fully represent the views and experiences some students in similar programs elsewhere. Secondly, it did not include clinicians’ perspectives on clinical supervision, which could have offered valuable insights into their roles and challenges. Therefore, further research involving multiple institutions and incorporating both student and clinician perspectives is recommended to better understand the impact of clinical learning experiences.

## Conclusion

This study examined nursing and midwifery students’ perceptions and experiences related to clinical learning and practice. Despite having a supportive clinical learning environment, students’ clinical learning was affected by limited engagement, perceived bias and negative attitudes from clinicians and inadequate clinical resources. Strengthening collaboration between training institutions and hospitals is essential to improve clinical preceptorship, enhance the quality of supervision, and better align educational curricula with practical training needs. Furthermore, investing in capacity-building initiatives and ensuring adequate resource allocation for clinical supervisors can significantly improve clinical learning outcomes.

## Data Availability

All data produced in the present study are available upon reasonable request to the authors.

## Acknowledgements

The authors would like to express their gratitude to the School of Medicine and Health Sciences, University of Papua New Guinea. The authors are also grateful to the nursing and midwifery students who participated in this research. Special acknowledgement to Dr. Carolyn Hastie from Griffith University, whose constructive feedback and suggestions significantly enhanced the quality of this manuscript.

